# Detecting cryptic clinically-relevant structural variation in exome sequencing data increases diagnostic yield for developmental disorders

**DOI:** 10.1101/2020.10.02.20194241

**Authors:** Eugene J. Gardner, Alejandro Sifrim, Sarah J. Lindsay, Elena Prigmore, Diana Rajan, Petr Danecek, Giuseppe Gallone, Ruth Y. Eberhardt, Hilary C. Martin, Caroline F. Wright, David R. FitzPatrick, Helen V. Firth, Matthew E. Hurles

## Abstract

Structural Variation (SV) describes a broad class of genetic variation greater than 50bps in size. SVs can cause a wide range of genetic diseases and are prevalent in rare developmental disorders (DD). Patients presenting with DD are often referred for diagnostic testing with chromosomal microarrays (CMA) to identify large copy-number variants (CNVs) and/or with single gene, gene-panel, or exome sequencing (ES) to identify single nucleotide variants, small insertions/deletions, and CNVs. However, patients with pathogenic SVs undetectable by conventional analysis often remain undiagnosed. Consequently, we have developed the novel tool ‘InDelible’, which interrogates short-read sequencing data for split-read clusters characteristic of SV breakpoints. We applied InDelible to 13,438 probands with severe DD recruited as part of the Deciphering Developmental Disorders (DDD) study and discovered 64 rare, damaging variants in genes previously associated with DD missed by standard SNV, InDel or CNV discovery approaches. Clinical review of these 64 variants determined that about half (30/64) were plausibly pathogenic. InDelible was particularly effective at ascertaining variants between 21-500 bps in size, and increased the total number of potentially pathogenic variants identified by DDD in this size range by 42.3%. Of particular interest were seven confirmed *de novo* variants in *MECP2* which represent 35.0% of all *de novo* protein truncating variants in *MECP2* among DDD patients. InDelible provides a framework for the discovery of pathogenic SVs that are likely missed by standard analytical workflows and has the potential to improve the diagnostic yield of ES across a broad range of genetic diseases.

## Main Text

Structural Variation (SV) includes a diverse collection of genomic rearrangements such as copy number variation (CNV), mobile element insertions (MEIs), inversions, translocations, and others^1^. Depending on population ancestry and technology used, the typical human genome harbours between 7,000-25,000 polymorphic SVs, with the majority constituting biallelic CNVs and MEIs^2^. While most SVs have minimal, if any, functional impact, SVs have been recognized as causative variants in congenital disorders^3–5^.

In diagnostic testing of suspected genetic disorders, SVs are often identified using chromosomal microarrays (CMAs) which offer a low-cost albeit low-resolution method for the identification of large CNVs (typically >20kbp in length for genic regions). CMAs are still widely used by diagnostic laboratories despite the increasing maturity of genome sequencing-based tools for SV discovery^6^ and the wealth of clinically-ascertained exome sequencing (ES) data already generated for the ascertainment of single nucleotide variants (SNVs) and small insertions/deletions (InDels)^7^. There are several reasons for this. First, the cost, computational power, and informatics complexity necessary for genome sequencing-based diagnostics is still a barrier to many public and private healthcare providers^8^. Second, current ES-based SV-discovery approaches focus on methods that interrogate sequencing coverage to identify regions of copy number variation within one genome compared to others^9^. As such, ascertainment is typically limited to CNVs of size >10kb, with resolution largely a factor of the sequencing depth and the density and number of baits in the ES assay, analogous to probes in CMAs. Thus, despite potentially offering improvements in CNV ascertainment over CMAs, ES as a tool for the assessment of diagnostic SVs has been slow to enter the clinic^10^.

Consequently, patients with genetic abnormalities smaller than the discovery resolution of CMA or standard SV-ES approaches (>10kb) but larger than variants able to be accurately called using typical SNV/InDel based approaches (<50bp)^11^ often remain undetected, here termed “cryptic”. To address this unmet need, we have developed the tool InDelible, which examines ES data for split read pairs indicative of SV breakpoints. We decided to focus on split reads because the formation of unique junction sequences is a shared characteristic of a broad range of different classes of SVs. We applied InDelible to ES data generated from 13,438 probands with severe developmental disorders (DD) recruited as part of the Deciphering Developmental Disorders (DDD) study. Approximately 29% of DDD probands harbour a pathogenic *de novo* mutation in a gene known to be associated with DD^7^ and have been previously assessed for a wide range of variant classes such as coding^7^, noncoding^12^, and splice site^13^ SNVs and InDels, multinucleotide variants^14^, mobile element insertions^3^ and copy number variants (unpublished). As such, the DDD study represents an ideal opportunity to demonstrate the additive diagnostic potential of identification of SVs at scale using split-read information.

InDelible variant discovery and analysis proceeds in several steps (Figure 1; detailed description in Supplementary Methods). In summary, Indelible identifies split reads, aggregates them into clusters at the same genomic location, filters these clusters to remove technical artefacts and retain likely genetic variants, and then combines unaligned portions of split reads and maps them to the genome to characterise the nature of the variant. InDelible also calculates the frequency of each split-read cluster across a population of individuals to facilitate the filtering of variants on the basis of minor allele frequency.

**Figure 1:**
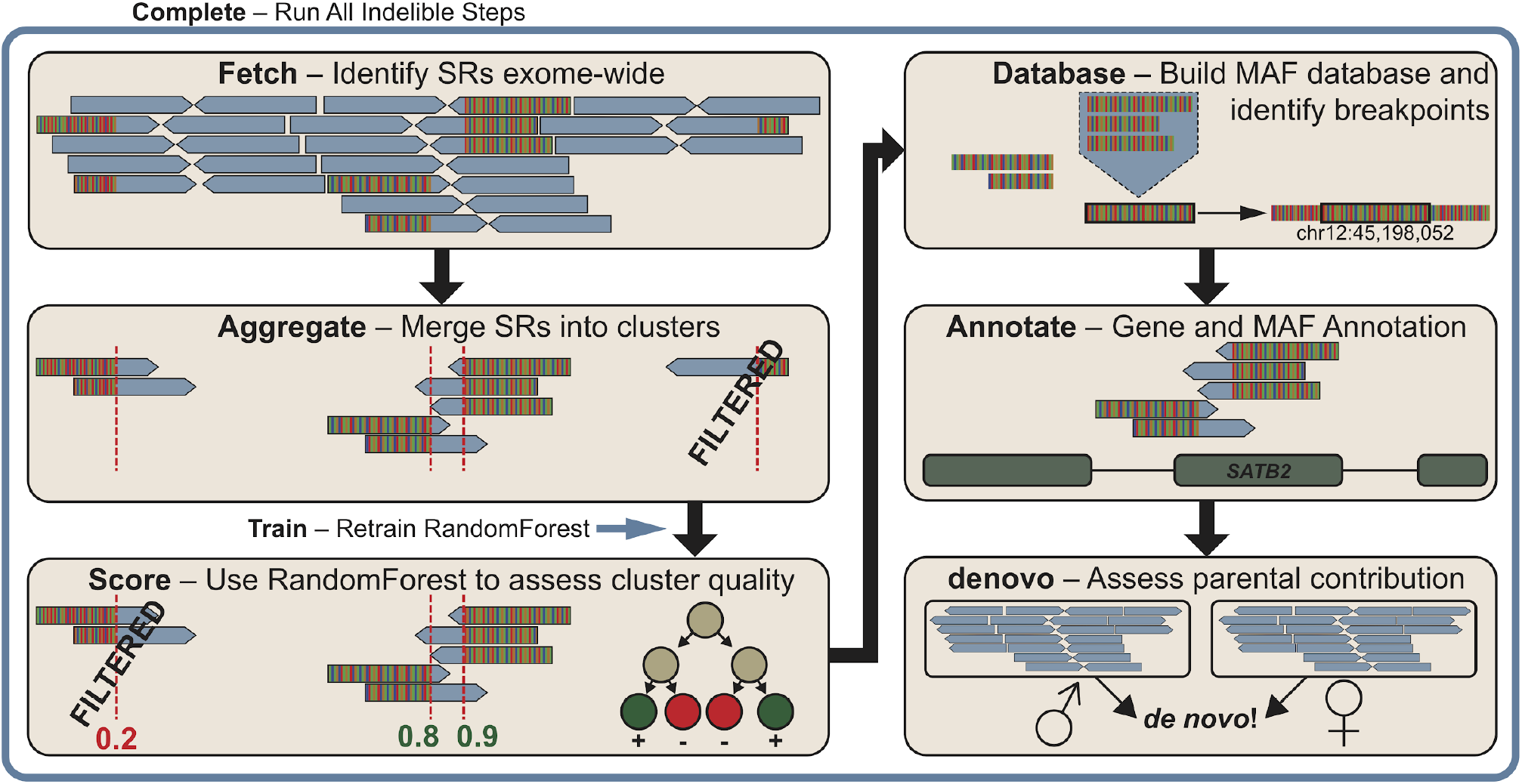
InDelible SV discovery in ES data. InDelible processes one ES sample provided in BAM or CRAM format via six primary steps (tan boxes). First, alignment files are queried for all reads where part of the aligned sequence matches the reference genome and the other does not (i.e. split reads; Fetch). Next, reads are clustered (Aggregate) and scored using a random forest model^15^ trained using a variant truth set (Score; see Supplementary Figure 1 and Supplementary Methods for more detail). Split reads are then merged within clusters across individuals to determine the longest quality junction sequence and mapped back to the genome with bwa mem^16^ and to a set of curated repeats with blastn^17^. These alignments are then used to determine breakpoint frequency, likely breakpoints, length, and structural variant class (i.e. deletion, duplication, insertion, etc.; Database). Split read clusters are subsequently annotated with population frequency and intersection with genomic functional annotations, such as protein-coding genes (Annotate). Finally, clusters are assessed for presence or absence in parental samples, where available, to determine inheritance status and identify likely de novo variants (denovo). All of these commands can be run on one sample via the “Complete” command (blue box). InDelible also includes the “Train” command to train a new random forest model from user-provided labelled training data.

InDelible is coded in Python, uses the pysam^18^ library for sequence alignment file manipulation (Supplementary Table 1), and works on bwa-aligned BAM or CRAM format files^19^. We have designed InDelible to be scalable for datasets comprising individual probands to multi-thousand sample cohorts and our estimates suggest that, to analyse a dataset of 1,000 trios, InDelible would require approximately 1544 CPU hours, or 15.4 hours of real time on a 100-core compute cluster (Supplementary Figure 2). Additionally, for easy implementation on cloud compute platforms, we have made InDelible available as a Docker image (see Supplementary Methods and Data and Code Availability).

We then benchmarked InDelible against GATK^11^ and Manta^20^, another SV detector which utilises split reads, for variants across a range of allele frequencies and sizes in a control individual from the gold-standard variant dataset generated by the Genome in a Bottle Consortium (Supplementary Figure 3; Supplementary Methods)^21,22^. When using ES data, InDelible equals or exceeds the recall of both GATK and Manta for variants between 21-10kbp in length, the variant space InDelible was targeted to identify. Relative to InDelible, GATK and Manta had 81.7% and 15.0% recall for deletions >20bp in length, respectively, and 86.9% and 8.2% recall for insertions >20bp in length, respectively. In this same experiment InDelible has moderately increased false discovery rates compared to GATK (Supplementary Figure 3). These issues can likely be attributed to InDelible being designed for maximum sensitivity in clinical sequencing data and can likely be abrogated via the design of better hard filters when analysing population-level variants and/or retraining the random forest using training data from population-level datasets.

A key objective for the design of InDelible was to identify *de novo* variants potentially causative of a proband’s disorder. As such, variants are primarily filtered on: (i) the population frequency of the split read cluster to remove variants too common to be plausibly causative of a rare disorder, (ii) absence in unaffected parents (when available), and (iii) intersection of variant breakpoints with the coding sequences of known-disease associated genes. Defining the precise molecular structure of SVs from short read sequencing data can be challenging, and even minor errors in breakpoint precision can have large consequences on interpretation (e.g. in versus out of frame InDels). Hence, we opted to identify all variants which intersect relevant DD-associated genes for further manual curation rather than relying on generic variant interpretation tools.

To evaluate the utility of InDelible for diagnostic analyses, we applied InDelible to identify putatively diagnostic variants in 13,438 probands recruited to the DDD study. Probands were exome sequenced either with both parents (trios, n = 9,848) or with one or both parents absent (non-trios, n = 3,590). We first identified split reads and split read clusters (Figure 1) to ascertain 353,313,108 redundant split read clusters across all probands. Random forest filtering resulted in retention of 30,667,420 high-quality, redundant split read clusters across all probands, or 8.7% of originally ascertained loci (Supplementary Methods, Supplementary Figure 4). After cluster filtering we merged all retained clusters into a set of 1,954,642 non-redundant split read clusters across all 13,438 probands, with 1,342,050 (68.7%) clusters found only in one proband (Supplementary Figure 5). Clusters were evenly distributed across all chromosomes as a function of chromosome length (r^2^ = 0.739; Supplementary Figure 6). Retained clusters were then annotated with putative breakpoints, intersecting gene(s), and population frequency. InDelible was also able to determine the missing 5’ or 3’ breakpoint, variant length, and variant type (i.e. deletion, duplication, MEI, etc.) of 199,932 (10%) clusters (Supplementary Methods). Of the clusters which InDelible was able to resolve to a specific variant type, 65.7% were simple deletions/duplications, with the remainder comprising complex events, MEIs, translocations/segmental duplications, and non-templated insertions (Supplementary Figure 7). Ascertainment of variant type and length are dependent on sequencing depth and population frequency (Supplementary Figure 5), but are optimized for the length of variants InDelible is best suited to identify (∼20-500bp; Supplementary Figure 8). This specificity is best demonstrated when restricting to clusters that are plausibly associated with patient phenotype (see below); InDelible accurately resolves both breakpoints, length, and variant type for 86.4% (127/147) of such clusters (Supplementary Data 1).

We next restricted our variant set to rare (call frequency <0.04%) clusters found only in or near (here defined as within ±10bp of any exon) the coding sequence of 399 dominant or X-linked DD-associated genes from the Developmental Disorders Genotype-to-Phenotype database (DDG2P)^23^. Variants identified within individuals sequenced as a parent-offspring trio were then also assessed for *de novo* status. Filtering on allele frequency, inheritance, and gene intersection resulted in a preliminary set of 260 candidate InDels and SVs across all 13,438 probands (Figure 2A; Supplementary Data 1; Supplementary Methods). Based on manual variant inspection^24^, we determined that 2/260 (0.8%) were erroneously annotated to have intersected a monoallelic DD gene, 17/260 (6.5%) candidate *de novo* events were likely to be present in a parent (i.e. parental false negatives), and 23/260 (8.8%) were unlikely to be real variants (i.e. offspring false positives). Four probands contributed 52.2% of false positive variants, indicating that sample selection and/or additional sample-level QC could further lower the false positive rate of InDelible (Figure 2A; Supplementary Data 1).

**Figure 2:**
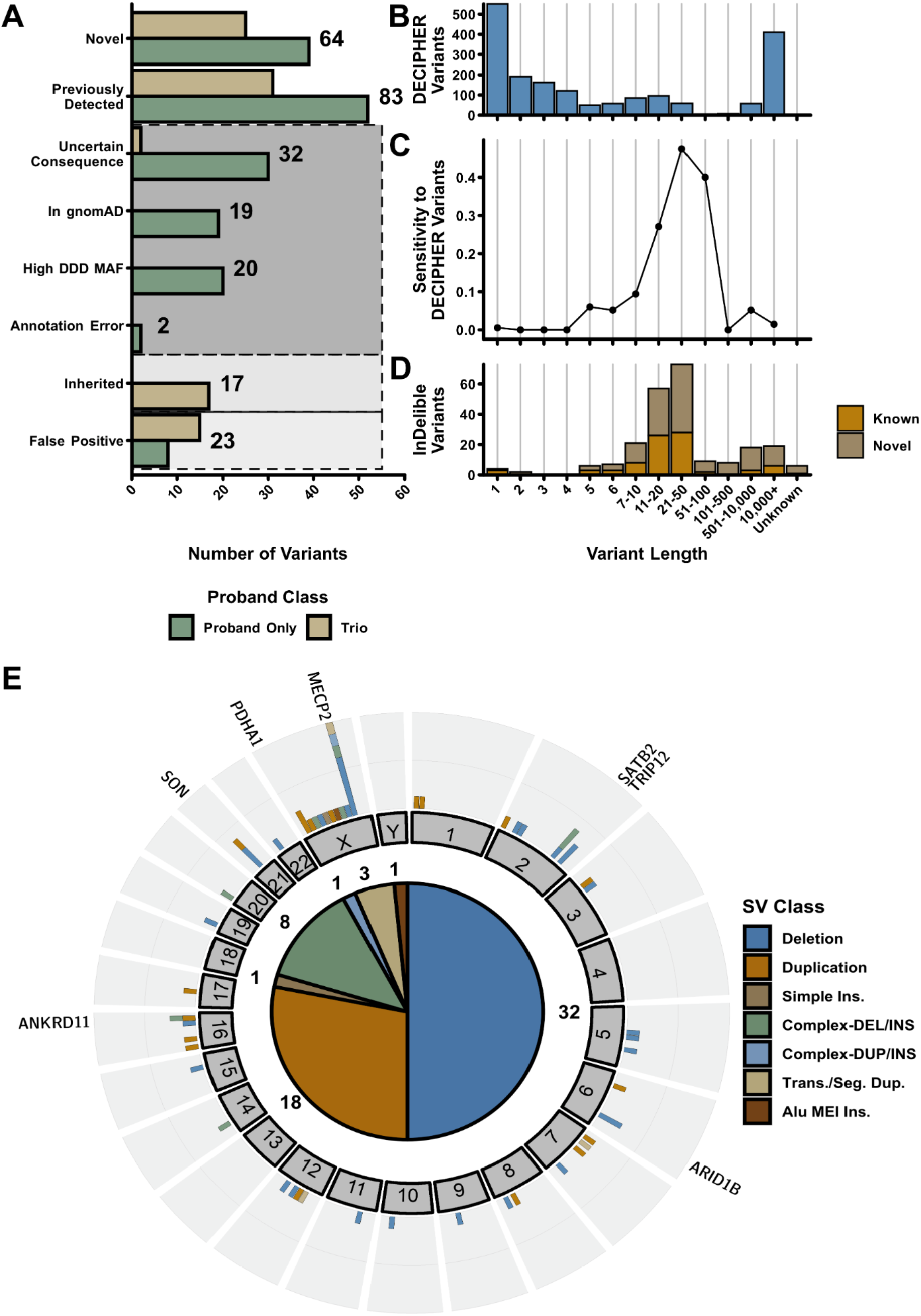
SV ascertainment in the DDD study with InDelible. (A) Breakdown of putative variant consequences for all 260 variants identified in this study delineated by whether or not the proband was sequenced with both parents (Trio – Tan) or not (Proband Only – Dark Green). Grey boxes represent erroneous variants and variants unlikely to be associated with a proband phenotype. (B) Total number of previously reported variants among DDD probands with a net size change ≥ 1bp. (C) Sensitivity of InDelible to DDD reported variants among various variant size bins. (D) Categorization of InDelible ascertained variants into previously known (Orange) versus those novel (Brown) to this study based on size. (E) Distribution of variants unique to InDelible throughout the genome. Shown in the outer plot are the total number of InDelible variants per gene, with genes which have multiple previously undetected variants labeled. Displayed In the inner plot are the total number of variants for each SV type identified.

Following variant quality control, we further curated variants for those likely to be associated with a proband phenotype (Figure 2A). We considered variants with a non-Finnish European minor allele frequency of ≥ 1×10^−4^ (19/260; 7.3%) in the genome aggregation database (gnomAD)^1,25^ or presence in other unrelated individuals within DDD (20/260; 7.7%) as unlikely to be the cause of the child’s disorder. Additionally, variants confined to introns or 5’/3’ UTRs were also defined as variants of uncertain significance and were not considered further (32/260; 12.3%). This final round of filtering left 147 SVs and large InDels which could plausibly explain a proband phenotype (56 from probands sequenced as trios, 91 from non-trio probands).

We next sought to determine the sensitivity of InDelible to clinically relevant variants ascertained using alternative methods. DDD has already identified (across both trio and non-trio probands) 1854 rare, plausibly pathogenic variants with a net size difference ≥ 1bp (i.e. non-SNVs) in the same DDG2P gene set defined above^7^ – variants potentially detectable with a split read-based method such as that employed by InDelible. The majority of these variants are private or low allele frequency small InDels between 1-10bp in size (1218/1854; 65.7%) or large CMA or ES-ascertained CNVs ≥ 10kb in length (410/1854; 22.1%; Figure 2B). As anticipated due to the low number of split reads at variant breakpoints as variant size decreases, InDelible performed poorly in identification of very short variants ≤10bp with an overall sensitivity of 1.4% (Figure 2C). Sensitivity improved as a function of variant size, peaking at 47.5% sensitivity for variants between 51-100bp, but dropped again for variants ≥ 100bp. To better understand why InDelible missed such variants, we manually curated the 34 potentially pathogenic variants between 20-500 bps not reported by InDelible. We found that InDelible missed variants for three primary reasons. First, these potentially pathogenic variants include some higher frequency variants that are too common to be plausibly pathogenic, whose true allele frequency was underestimated previously, but has been more accurately determined by Indelible, and then subsequently filtered out (n = 12/34; 35.3%). Second, several variants have low split read support (i.e. <5 reads) despite being located in high coverage regions and were thus excluded by our stringent filtering approach (n = 11/34; 32.4%). Third, as variant size increases, it becomes more likely that the breakpoints of SVs which impact coding sequence lie outside of ES target regions (i.e. within intronic and intergenic sequences). Ergo, such variants are refractory to identification with split reads and likely to be missed by any split-read caller (n = 6/34; 17.6%). Combined, these three explanations account for 85.3% of variants between 20-500bps missed by InDelible. While variants with breakpoints outside of sequencing baits are invisible to InDelible, additional fine-tuning of InDelible’s filtering parameters could, in theory, output variants with lower split read support or variants with higher allele frequencies.

These 64 previously undetected variants (four of which were included as part of a previous DDD publication^26^) that impact known DD-associated genes (Supplementary Data 1) are composed primarily of deletions and duplications (50/64; 78.1%) but also includes variants with diverse mutational mechanisms such as MEIs, complex rearrangements, and dispersed duplications/translocations (Figure 2E). Twenty five of these variants were observed in trio probands, with parental data supporting a *de novo* origin for all of these variants. InDelible was particularly effective at identifying variants between 21-500bps in size (Figure 2D) – 30 previously undetected variants (46.9% of InDelible-specific variants) lie within this size range and represent a 42.3% increase in putatively pathogenic variants 21-500bps in length among DDD probands (Figure 2D). We also identified four genes with multiple previously undetected SVs among unrelated individuals, of which the most recurrently affected was *MECP2*, the causal gene of Rett Syndrome (Figure 2E)^27^.

From an initial round of clinical review, based on intersecting gene(s) and associated phenotypes, we concluded that ten (15.6%) of these 64 previously undetected variants were unlikely to explain the referred proband’s phenotype, and were thus excluded from future analysis (Supplementary Data 1). We next attempted PCR validation of the 54 putatively pathogenic variants (Supplementary Methods). Of the variants for which conclusive validation results could be obtained, 23/23 (100%) were confirmed as true positives, either by the obvious presence of a mutant band of expected size with gel electrophoresis or by follow-up capillary sequencing where the gel result was uncertain (Supplementary Data 1). For variants for which PCR was possible, we also confirmed that 10/10 (100%) putative *de novo* variants identified in trio probands were indeed absent from both parents.

All 54 plausibly pathogenic variants were then clinically interpreted by two senior clinical geneticists; 30/54 (55.6%) were classified as pathogenic or likely pathogenic by both clinical geneticists (Supplementary Data 1). Of these variants, those identified in non-trio probands (n = 30/54 plausibly pathogenic variants) for which inheritance status is unavailable, were less likely to be interpreted as being pathogenic (Fisher’s p = 0.016). This finding is corroborated by the difference in the proportion of in-frame versus out-of-frame deletions and duplications ≤50bp between trio and non-trio probands; 78.2% of deletions and duplications are in-frame for non-trios versus 19.0% for trios (Fisher’s p = 3.4×10^−6^; Supplementary Figure 9). This is consistent with population-level observations – out-of-frame deletions and duplications are typically under stronger negative selection than in-frame variants^28^ and an increased proportion of in-frame variants in non-trio probands is suggestive of a greater proportion being benign. The difference is likely attributable to the absence of parental data leading to the inclusion of rare benign inherited variants that are unlikely to be filtered out using population variation data (e.g. gnomAD^1,25^). Overall, *de novo* variants identified by InDelible represent 0.7% (18/2592) of all confirmed diagnoses among trio probands in the DDD study.

InDelible identified a total of seven confirmed *de novo* variants ≥ 20bp in length affecting *MECP2* (Figure 2E; Figure 3A), all predicted to be protein truncating. As expected and in accordance with known sex bias among Rett Syndrome patients^29^, all variants were ascertained from female probands. Out of these seven probands, two have phenotypes that could be described as consistent with typical Rett Syndrome presentation^29^. Through in-depth clinical curation of HPO terms (see Supplementary Methods), we grouped probands with putative loss of function mutations caused by SVs in *MECP2* into four categories (Figure 3B). Cases identified by InDelible thus represent the wide variety of diverse clinical presentations that can result from disruption of the C-terminus of *MECP2*^*30*^ and include previously observed *MECP2*-associated phenotypes such as early onset seizures and Angelman-like symptoms (Supplementary Data 2; Figure 3B)^31^.

**Figure 3:**
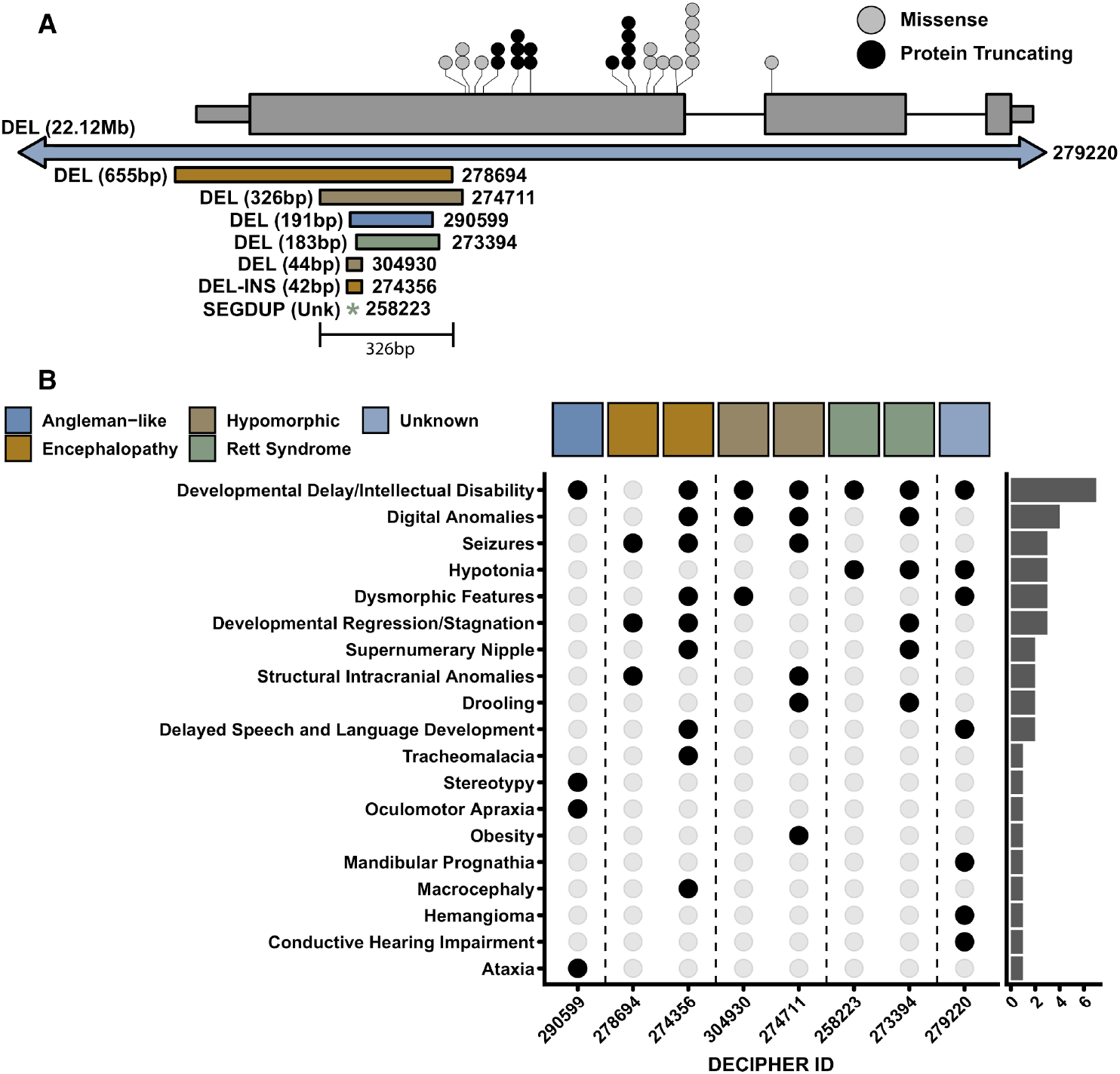
Clustered SVs in MECP2 cause diverse phenotypes. (**A**) Shown is a cartoon representation of the gene MECP2, with stop-gained (black circles) and missense (grey circles) de novo SNVs identified in DDD trios. Each circle represents one proband, with recurrent variants represented by stacks of circles. Below the MECP2 gene model, we have shown the seven variants identified by InDelible as well as the single whole gene deletion previously identified via CMA (proband 279220; arrows indicate this variant extends beyond the scale shown in the diagram). Sizes adjacent to variants represent the total number of alternate (DEL + INS) bases attributable to each variant. We have indicated that the variant in patient 258223 only incorporates non-references bases (i.e. an insertion) with a ‘*’. All InDelible-ascertained variants overlap the same 326bp region in the last exon of MECP2. (**B**) Diverse proband phenotypes among MECP2 SV carriers. Each proband carrying a MECP2 SV from (A) is shown on the x-axis, with phenotypes annotated by the referring clinician shown on the y-axis. Filled black circles represent when a corresponding proband displays the corresponding phenotype. Colored boxes on the top of the plot represent the diverse phenotypes we identified following clinical review. The y-axis marginal histogram represents the number of times the corresponding phenotype was observed among our SV probands.

Interestingly, all five of our *MECP2* variants in probands without typical Rett syndrome presentation overlapped the same 326bp region located within the final coding exon and, aside from a previously ascertained whole gene deletion (proband 279220), do not overlap with putatively pathogenic SNVs identified within the DDD study (Figure 3A). The SV-specific region corresponds to an area of low sequence complexity and has been previously ascertained as hyper-mutable by several studies^30,32^. The molecular function of this region of *MECP2* is poorly understood and it is uncertain as to the consequences that our described variants may have on protein structure beyond decreasing transcript abundance and/or overall protein stability^30^.

The seven *de novo MECP2* variants constitute 28.0% (7/25) of all novel *de novo* variants identified by InDelible, and 35.0% (7/20) of all confirmed *de novo* protein-truncating or gene-deleting variants of *MECP2* in the DDD study^7^ (Figure 3A).

As several publications have shown that rare, inherited variants are also important in the genetic architecture of developmental disorders^33^, we next sought to examine if InDelible could be used to identify such variants. We repeated our filtering as described above, but limited to variants found in only a single proband with split read support from either parent (Supplementary Methods). This approach identified a total of 145 variants within the coding sequence of monoallelic DD genes. As expected based on our analysis of variants in probands sequenced without their parents (Supplementary Figure 9), a large proportion of inherited variants we identified were balanced/in-frame deletions or duplications with uncertain effect on the target protein (50; 34.5%). Others either primarily overlapped noncoding sequence, were found in an individual with a more likely diagnostic variant, were large duplications which only partially overlapped the gene of interest, were already identified based on an alternate breakpoint as part of our *de novo* analysis, or were also identified in control individuals at high enough allele frequencies to be considered unlikely to be associated with patient phenotype^25^. Initial filtering based on these criteria left a remainder of 17 variants for clinical interpretation.

Of the remaining inherited variants, 7 were already identified via other approaches and reported to referring clinicians with 6 considered as likely benign and 1 as likely pathogenic. The remaining 10 variants were referred to the same two senior clinician geneticists as for our *de novo* analysis detailed above (Supplementary Data 3). Of these 10 variants, all but 1 were unlikely to be involved in patient phenotype. The sole remaining inherited variant, an out-of-frame deletion in *KAT6B*, was identified in a proband-mother pair. This variant was deemed a variant of uncertain consequence upon initial clinical review. Follow-up with the referring clinician regarding the mother’s phenotype determined that the mother did not exhibit any features of the proband’s disorder. As such, this variant was deemed to be likely benign. Combined, this data shows that InDelible is effective at identifying rare, inherited variants but that the overall diagnostic yield may be low.

Here we present the development and application of InDelible, a novel tool designed for the rapid assessment of ES data for breakpoints of rare, pathogenic cryptic SVs involved in single gene disorders (Figure 1). We applied InDelible to 13,438 proband genomes sequenced as part of the DDD study and identified a total of 147 candidate pathogenic variants impacting genes associated with dominant or X-linked DD (Figure 2A, Supplementary Data 1). Of these 147 variants, 64 were not previously identified in DDD probands, despite the wide range of SV and InDel detection algorithms that have previously been deployed on this cohort^7,26^. Notably, we increased the number of putatively diagnostic variants among DDD probands 21-500bp in length by 42.3% (Figure 2D). Through conservative clinical assessment of these 64 variants, we determined that 30 (46.9%) of our previously undetected variants were considered likely causative of proband phenotype – of particular interest was the large number of protein truncating SVs we identified in *MECP2* (Figure 3).

The variant size range which InDelible interrogates is complementary to other approaches commonly used for variant discovery from ES data^9,11^. While other previously described algorithms have also attempted to mine split read information for structural variant detection^11,20,34^, they have different properties that preclude meaningful comparison with InDelible^11^. Some have been trained primarily on genome sequencing data rather than ES data^20,34^, others do not explicitly assess *de novo* status, and many are not readily scalable to a dataset of ∼10,000 trios. As such, we have built InDelible to be scalable to many thousands of samples (Supplementary Figure 2).

Other studies have previously noted that ∼10% of all *MECP2* variants in probands ascertained based on presentation of Rett-associated phenotypes were deletions^32,35^ and a large number of pathogenic or likely pathogenic variants in ClinVar fall within the same region of *MECP2* that we report in this manuscript. These observations, combined with the diverse phenotypes that this study has identified (Figure 3B), further complicate the clinical interpretation of variants disrupting *MECP2*. In particular, the work of Guy et al.^30^ found that slight differences between the size and sequence context of deletions in the C-terminal domain of *MECP2* can have significant ramifications in RNA/protein expression. Additionally, Huppke et al.^36^ found that skewed X-inactivation could play a role in the severity of *MECP2* presentation. Further work is needed to understand how different classes of mutation lead to diverse phenotypes in patients with *MECP2* loss of function variants. However, most importantly and exemplifying the additive power of InDelible, if not applied to the DDD study, 20.6% of DDD probands with clinically relevant *MECP2* variants would not have received a diagnosis for their disorder.

InDelible was designed to detect variant breakpoints missed by other approaches in ES data from DD patients. This has three major ramifications for the design of InDelible and the variants reported as part of this study. Firstly, as the primary cause of DD are highly penetrant dominant *de novo* variants^7^, InDelible variant reporting was focused on identifying such variants from a defined list of genes known to be associated with DD^23^. As briefly demonstrated above for rare inherited variation, this does not preclude the use of InDelible to identify variants acting through other modes of inheritance – InDelible will identify variants across the entire allele frequency spectrum and outside of the provided gene list as part of the primary output.

Secondly, the DDD cohort has been previously investigated for a broader range of variant classes (using both different assays and algorithms) than most ES studies. For ES-based CNV discovery from read-depth, DDD applied four separate algorithms to build a joint call set (unpublished). Thus, the added diagnostic value of running InDelible is probably under-estimated in the DDD study compared to other ES studies and/or common clinical sequencing practices which would be unlikely to utilize complex joint-calling approaches such as our own. To quantify the added diagnostic value of running InDelible across different settings by a user seeking to run a minimal number of algorithms, we estimated the proportion of unique PTVs InDelible would find if used alone or jointly with other algorithms targeting a breadth of variant types (SNVs, InDels, large deletions, and MEIs; Supplementary Methods)^3,9,11^. Overall, and when using other approaches, InDelible-specific variants will likely represent between 2-3% of all PTVs in a given cohort (Figure 4). This observation strongly implies that workflows that do not incorporate algorithms capable of detecting this class of cryptic variation are only likely to achieve 97-98% sensitivity for pathogenic PTVs.

**Figure 4:**
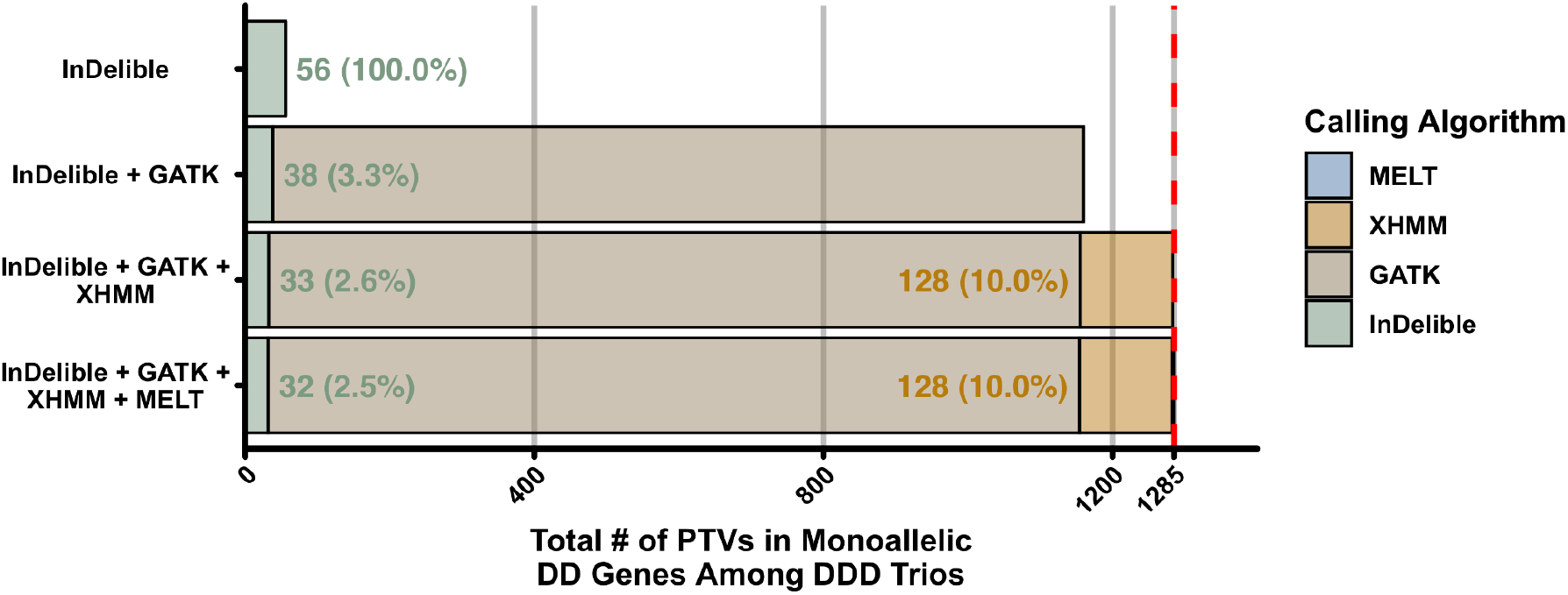
Added diagnostic PTV Yield of InDelible. Total number of de novo PTVs (y-axis) ascertained in DD-associated genes when using InDelible alone, or in combination with a subset of three additional algorithms (GATK^11^, XHMM^9^, or MELT^3,37^). Percentages represent the proportion of all PTVs specific to InDelible (green text) or XHMM (orange text) for each bar. The red line and axis label indicates the maximum number of de novo PTVs identified in DD-associated genes among 9,848 DDD trio probands if combining data from all four algorithms (n = 1,285 variants).

Finally, we note that InDelible is unlikely to be more effective than currently available tools when applied to genome sequencing data. In ES, discordant read pairs are typically much less informative for detecting SVs than in genome sequencing due to the inherent properties of the data. In genome sequencing, data combining split and discordant read-pair information is a better means to identify most SV types.

InDelible provides a rapid framework for the assessment of ES data for intermediate length pathogenic SVs of diverse mutational origins. Our results show that through a combination of improved algorithm design, variant annotation, and clinical interpretation, ongoing interrogation of well-studied datasets will continue to yield novel diagnoses.

## Data Availability

Sequencing, phenotype data, and variant calls for all data in this paper are accessible via the European Genome-phenome Archive (EGA) under study number EGAS00001000775 [https://www.ebi.ac.uk/ega/studies/EGAS00001000775 [ebi.ac.uk]]. Within this study, WES files of all DDD families are provided as part of the dataset EGAD00001004390. Gene lists and input data for analysis are available as part of the InDelible software.

https://github.com/HurlesGroupSanger/indelible

## Acknowledgements

We would like to thank the DDD participants and their families – without their trust and confidence this work would not be possible. We also wish to acknowledge Jeffrey Barrett for his leadership role in the DDD Study. The research team acknowledges the support of the National Institute for Health Research, through the Comprehensive Clinical Research Network. Informed and written consent was obtained for all families and the study was approved by the UK Research Ethics Committee (10/H0305/83, granted by the Cambridge South REC, and GEN/284/12 granted by the Republic of Ireland REC). The DDD study presents independent research commissioned by the Health Innovation Challenge Fund [grant number HICF-1009-003], a parallel funding partnership between Wellcome and the Department of Health, and the Wellcome Sanger Institute [grant number WT098051]. The views expressed in this publication are those of the author(s) and not necessarily those of Wellcome or the Department of Health. This study makes use of DECIPHER [http://decipher.sanger.ac.uk [decipher.sanger.ac.uk]]), which is funded by the Wellcome.

## Data and Code Availability

Sequencing, phenotype data, and variant calls for all data in this paper are accessible via the European Genome-phenome Archive (EGA) under study EGAS00001000775. InDelible is available at the InDelible GitHub repository: https://github.com/HurlesGroupSanger/indelible. All code and data used to generate figures and results in this manuscript is located at the following GitHub repository: https://github.com/HurlesGroupSanger/indelible_paper. A Docker image of InDelible is available at the following GitHub repository: https://github.com/wtsi-hgi/indelible-docker/tree/master.

## Author Information

A.S. performed initial design and implementation of the InDelible algorithm. E.J.G. provided additional code, prepared InDelible for publication, assessed the DDD cohort for variation with InDelible, and together with A.S. and M.E.H. designed experiments, oversaw the study, and wrote the manuscript. S.J.L, E.P., and D.R. designed and performed PCR validation experiments of InDelible-ascertained variants. G.G. and R.Y.E curated and maintained DDD sequencing data. P.D. curated ES CNV calls. H.C.M. helped curate DECIPHER variants and DDD patient diagnoses. C.F.W, D.R.F, and H.V.F performed clinical interpretation and helped write the manuscript.

