## Supplementary Materials for "Detecting cryptic clinically-relevant structural variation in exome sequencing data increases diagnostic yield for developmental disorders"

---

[*] Contributed equally

[a] Present address: Medical Research Council (MRC) Epidemiology Unit, University of Cambridge School of Clinical Medicine, Institute of Metabolic Science, Cambridge Biomedical Campus, Cambridge, UK

[b]

### Supplementary Methods

#### Overview of the InDelible Algorithm

InDelible is open source software licensed under GPLv3.0 and developed in version 3.7 of the Python programming language. InDelible is freely available for academic use and detailed installation and usage instructions are available at GitHub (see main text Data and Code Availability). While all results in this manuscript use the version of InDelible built on Python3.7, InDelible has also been tested for backwards compatibility with Python2.7. InDelible additionally makes use of several open source libraries, all of which are listed in Supplementary Table 1, and supports both CRAM and BAM input aligned with the bwa mem or aln algorithm[1].

A typical InDelible workflow proceeds in the steps outlined in main text Figure 1. Here we provide detailed descriptions of each of these steps:

- Fetch: Iterates through all reads in the provided sequence file and identifies those which have at least one end marked as clipped (i.e. with at least one "S" tag in the alignment cigar string) by bwa mem/aln[1]. Reads which, based on configurable user input, have a high map quality, split read length, and base quality are retained and provided as output for the next step.
- Aggregate: Reads which pass initial filtering in "Fetch" are then clustered based on chromosome and position. Clusters with fewer than a user-provided number of reads (default <3 reads), are discarded at this step. At each cluster, InDelible calculates features (Supplementary Figure 1) that will then be used as input to random forest (Supplementary Figure 4). At this step, InDelible also quantifies the proportion of reads at each cluster which have both the 5' and 3' end marked as split for later quality control (see "Detailed InDelible Variant Calling Protocol" below).
- Score: Each split read cluster is then annotated with the probability of being a true variant, regardless of clinical significance or *de novo* status, based on a 500-tree forest trained on manually curated and visualized truth data from DDD trios (see below)[2]. By default, clusters which have a probability <0.6 of being true sites are filtered at this step (Supplementary Figure 4).

- Database: Calculates allele frequency, longest alternate sequence, and SV class (i.e. deletion, duplication, insertion, etc.) from a provided set of files generated by the "Score" command. Calculation of frequency is done by simple overlap across all provided files, and does not consider nearby loci. For all merged loci, InDelible then attempts to find the longest split-read sequence across all merged individuals by identifying the longest sequence with at least 60% homology to all other sequences using the 'difflib' package for base Python. This sequence is then aligned to a set of curated human repeat sequences with the BLAST implementation included as part of biopython[3,4] and queried against the provided reference genome using bwa mem[1]. For bwa mem alignment, sequences are also paired with a simulated read extracted from up or downstream reference sequence to anchor the event. SV type, alternate sequence, breakpoints, and variant length are then extracted from bwa alignments. Only bwa alignments with mapping quality of both reads > 0, alignment orientation of both reads in the sense strand, and with the "reference" read anchored at the expected position are retained. While alignment orientation other than in the sense direction can be expected for a small number of variant types such as inversions, we opted to use this filtering regime to remove false positives. This filtering approach does not affect variants with at least one breakpoint directly impacting the coding sequence of genes included in the DDG2P target panel. The database step can also take a collection of priors; InDelible comes pre-packaged with a database generated from all probands analysed in this study.
- Annotate: Split read clusters are annotated with gene intersect to all ENSEMBL genes[5], disruption of coding sequence, and constraint based on the probability of being haploinsufficient[6]. A second gene list can also be provided which represents genes the end-user is specifically interested in (in the case of this manuscript, monoallelic Developmental Disorders Genotype-to-Phenotype database (DDG2P) genes[7]). Finally, the proportion of samples which also contain this cluster (i.e. allele frequency) is determined for each locus from the user-provided allele frequency database. All inputs at this stage can be modified by the end user to better fit specific project requirements.
- denovo: When provided, InDelible will query parental sequencing files for presence/absence of each split read cluster. Following assessment, clusters are then filtered if the total number of supporting split reads in either parental sample is too high (default >3 reads) and at sufficient coverage (default

≥9x). When parental samples are not available, “denovo” reformats the output from “Annotate” to make it consistent with output from probands sequenced as part of a trio. Columns requiring parental information are provided as “NA”.

Two additional commands are also provided with InDelible which add additional functionality depending on the needs of the user:

- Complete: Runs all of the above commands in succession for one sample.
- Train: Trains the random forest which InDelible uses to determine likelihood of being a true variant for each split read cluster. This method uses the random forest implementation provided as part of the Python package scikit-learn[2] combined with active learning. Active learning uses an iterative approach whereby a test and training set of size $k$ are constructed from a pool of $n$ variants of known truth with equal class balance, with all remaining variants (i.e. $n - k * 2$) placed into a validation set – the goal of active learning is to focus model training on the categorization of variants that are most difficult to classify. The random forest is then constructed with the training set and accuracy of the model determined via prediction on the test set using several covariates calculated during the ‘Aggregate’ step (Supplementary Figure 1). $k$ sites are then moved from the validation set to the training set based on their inability to be classified compared to other variants ( $|P(TP) - P(TN)|$). The random forest is then retrained using the new training set until the change in prediction accuracy of the test set between rounds is less than 1%. Random forest settings as implemented by scikit-learn are set to default except the number of computed decision trees, which is set to 500. To avoid issues with class balance, the Train command randomly samples the same number of false sites as true sites. InDelible includes training data comprising variants ascertained from the DDD study and the resulting random forest model, but can be retrained with alternative data.

As part of the InDelible distribution, when possible, we have provided genomic reference files such as gene models, DDG2P genes included in this study, and repeat sequences. Instructions to download files that are not provided (e.g. the human reference) are provided as part of our github repository. The output of the “Database” and “Train” commands generated with DDD data are also provided (see Data and Code

Availability in the main text), but in most cases it is highly recommended to regenerate these files with data specific to the current project.

We have also generated a Docker image containing the version of InDelible used to analyse the DDD data included in this manuscript. Functionality is identical to that described above. Please see the Data and Code Availability section of the main text for more information on acquisition, installation, and usage of the Dockerised version of InDelible.

#### Patient Recruitment and Sequencing

A total of 13,451 patients were recruited from 24 regional genetics services throughout the United Kingdom and Republic of Ireland as previously described[8]. Sequencing of families and alignment to the human reference genome (GRCh37) with bwa[1] was performed as previously described[8] but is repeated here in brief. Genomic DNA from all samples (probands and recruited parents, if available) recruited to DDD were fragmented to an average size of 150bp and used to create Illumina PCR-amplified paired-end libraries. Libraries were then hybridized to one of two SureSelect RNA baits (v3 or v5), captured, amplified and submitted for 75bp paired-end sequencing on an Illumina HiSeq following manufacturer instructions. All samples were sequenced to a mean depth of 90x across primary bait capture regions. Following sequencing, all samples were aligned with either bwa aln or mem[1] to version the 1000 Genomes Project phase 2 reference (vers. hs37d5) and processed with IndelRealigner and Base Quality Score Recalibration (BQSR) available as part of the GATK resource bundle (version 2.2).

#### InDelible Variant Calling Protocol

To ascertain the variants reported in this manuscript, we applied InDelible to all 13,451 recruited probands, but excluded 13 probands from further analysis due to excessive runtime, leaving 13,438 probands. This includes probands sequenced with both parents (trios, n = 9,848) or with one or both parents absent (non-trios, n = 3,590). As this was the first dataset analyzed with InDelible, we ran all steps while also training the random forest (Figure 1).

All probands were first run through the “Fetch” and “Aggregate” steps with default settings to identify 353,313,108 redundant split read clusters. Next, to train our random forest to perform split read cluster quality control, we randomly selected 2,000 non-redundant sites across all probands from the output of the aggregate step. We then visually inspected all 2,000 sites using the Integrative Genomics Viewer[9] to build a labelled truth set of variants for training. These 2,000 manually curated sites were then provided as input to the "Train" subcommand of InDelible at various test and training sizes ($k$). We ultimately decided on a probability ($p$) of being a true variant $p > 0.6$ at $k = 75$ as a reasonable value for filtering following cross-validation (Supplementary Figure 4). We then used the trained random forest to score all sites identified with the probability of being a true variant with the InDelible "Score" command. Training data used for this study is available as part of the GitHub repository provided in the Data and Code Availability section of the main text.

We next provided all output files of the "Score" command for all probands as input to the "Database" command with default settings to build the allele frequency database required as input to the "Annotate" command. All probands and split read clusters were subsequently processed with the "Annotate" and "denovo" commands with default settings. Following initial calling, we performed additional quality control to generate our final set of putatively clinically relevant variants. Split read clusters were retained at this step based on the following criteria:

1. Breakpoint frequency < $4 \times 10^{-4}$
2. Average MAPQ ≥ 20
3. Number of split reads ≥ 5
4. Proportion of split reads as a factor of coverage ≥ 0.1 (i.e. sr_total / coverage)
5. Found within coding sequence (here defined as exons ±10bp)
6. Intersected a gene with a known monoallelic, X-linked, or hemizygous mechanism with a loss of function or dominant negative consequence based off of the DDG2P database[10]
7. < 2 split reads in either the maternal or paternal sample if available
8. < 50% of reads with both ends split (i.e. both 5' and 3' ends with an "S" tag in the cigar string) AND
    a. ≤ 10% of reads with both ends split OR
    b. > 10% of reads with both ends split while also having a valid bwa alignment (see "Database" step)

A script which performs this filtering is included at the InDelible GitHub repository (see main text Data and Code Availability). Calling and subsequent filtering left a remainder of 354 split read clusters. Split read clusters identified at the same locus within the same proband were subsequently manually merged (marked as "OTHERSIDE" in Supplementary Data 1), leaving a remainder of 260 variants, with the 5' breakpoint retained for final reporting.

Via visual inspection of read alignments[9], we next determined whether variants were likely to be real in the proband and inherited from a parent where possible (Figure 2A). Variants with gnomAD non-Finnish European allele frequency ≥ $1x10^{-4}$ based on Karczewski et al.[11] (variants ≤ 50bp) or Collins et al.[12] (variants > 50bp) were then filtered out from further analysis (Figure 2A, Supplementary Data 1). This approach is imperfect – as the variant size range that InDelible detects is under-represented, some variants may be common in the population but may not be represented in these reference datasets. To determine the sensitivity of InDelible for DDD variants previously reported as potentially pathogenic, InDelible variants were intersected with previously reported DDD variants[10] and with CNVs called from read-depth analysis of DDD ES data (Supplementary Data 1). Previously known variants, false negative/positive variants, variants with high DDD/gnomAD allele frequency, variants that do not actually intersect a known DD gene, and variants located in regions difficult to interpret clinically (e.g. intron or 5'/3' UTR) are annotated as such in Supplementary Data 1.

To identify a set of rare inherited variants that could plausibly be associated with patient phenotype, we repeated our above filtering as for *de novo* variants except we restricted to variants found only in a single proband (e.g. singletons) without filtering for parental split read support. This approach identified a total of 211 breakpoints which, after collapsing identical loci as above, left a total of 145 variants for downstream analysis. We next excluded variants based on likely association with patient phenotype by removing:

- In-frame InDels inherited from an unaffected parent.
- Primarily non-coding variants.
- Variants found in patients with a more plausible variant already reported.
- Variant types of uncertain consequence such as processed pseudogenes and duplications which partially overlap coding sequence.

- Presence in any control individuals in the gnomAD database.
- Likely *de novo* variants already ascertained by InDelible and/or other approaches

This filtering left a total of 17 variants, of which 7 were already identified by an alternate approach and returned to referring clinicians. The remaining 10 variants were annotated for contribution to patient phenotype, returned to referring clinicians where relevant, and provided in Supplementary Data 3.

#### Benchmarking InDelible Runtime and Memory

To benchmark InDelible, we collected usage statistics provided by the standard output of the Platform Load Sharing Facility (LSF) at the Wellcome Sanger Institute. Metrics were generated during the course of processing the 13,438 DDD individuals through the standard InDelible SV discovery pipeline. The compute cluster used consists of 105 nodes with 32 8-core 2400Ghz CPU AMD Opteron Processors with 256Gb of memory each. To generate the extrapolated curve seen in Supplementary Figure 2C, we randomly sampled individuals 100 times at each sample size to generate an average expected runtime. On average, InDelible took 92.9 CPU minutes and a maximum of 1.2Gb of memory to run one sample with mean exome-wide coverage of 90x from aligned CRAM file to reporting of candidate diagnostic variants (Supplementary Figure 2). Considering that most users would likely be utilizing InDelible to analyse multi-sample datasets, we also calculated extrapolated runtimes via down-sampling of our own DDD runtimes for datasets composed of between 1-10,000 individuals (Supplementary Figure 2).

#### PCR Validation

To validate all 54 variants returned to clinicians via the DECIPHER platform, we used PCR. For small variants (≤50bp) we extracted the surrounding 300bp and used Primer3[13] with GC clamp turned on to automatically design an initial set of primers. Following initial design, we confirmed specificity with UCSC *in silico* PCR. For larger variants, we manually designed primers 5' and 3' of the predicted variant breakpoint and likewise confirmed specificity with UCSC *in silico* PCR. PCR was carried out using REDAccuTaq® LA DNA Polymerase (Sigma); 80ng of genomic DNA extracted from blood or saliva was amplified in the presence of 0.4 µM of each primer and 1 unit of REDAccuTaq. We used the following cycling conditions: 30s at 98°C followed by 6 cycles of (15s at 94°C, 45s at 60°, and 60s at 68°C) and 24 cycles of (15s at

94°C, 45s at 55°, and 60s at 68°C). For longer variants (i.e. ≥1Kbp) elongation time was changed from 45s to 480s. A final elongation step was carried out for 30m at 68°C. PCR products were visualized on a 2% or 4% agarose gel for short and long products, respectively. Following PCR, all reactions were PCR purified using a standard exosap protocol and submitted for capillary sequencing. Following sequencing, traces were examined for quality and BLAT and manual inspection used to check for presence of the predicted variant. For 28 variants we were unable to obtain conclusive validation results due to too little DNA for PCR (n = 3), failed capillary sequencing (n = 8), failed primer design (n = 6), or failed PCR (n = 11). We were unable to allocate resources during the COVID-19 pandemic to validate 3 variants prior to returning them to referring clinicians.

#### Analysis of *MECP2* Carrier Phenotypes

For proband phenotypes reported in Figure 3B, we first collated all HPO terms as reported by the referring clinician for all probands with an InDelible-ascertained *MECP2* variant. We then condensed these terms into 19 super-HPO groups which group related HPO terms into a more broad phenotype (e.g. the terms long toe and long fingers condense into the digital anomalies super-HPO group). Original HPO terms for all probands and the super-HPO terms to which they were assigned are available in Supplementary Data 2.

#### Proportion of PTVs Called By InDelible

To determine the proportion of PTVs within a dataset attributable to InDelible as shown in Figure 4, we queried variants identified in DDD probands sequenced as a trio from three different sources: (i) GATK called SNVs and short InDels ≤100bp from DECIPHER for DDD patients (n = 1,140 variants)[8], (ii) XHMM called CNVs from CNV data generated for DDD probands (unpublished; n = 128 deletions), and (iii) MELT[14] called MEIs from Gardner et al.[15] that were plausibly associated with patient symptoms (n = 4 MEIs). For all variant types, we retained only *de novo* variants which overlapped the same set of DD genes used for InDelible ascertainment. For InDelible, we considered the 56 *de novo* variants reported in Supplementary Data 1, regardless of novelty. InDelible Variants were then matched to each of the three other callsets, first by exact coordinate/sequence match and then by subsequent manual confirmation. Variants queried for the purpose of this exercise were also used to catalogue previously reported *MECP2* variants.

#### Benchmarking InDelible using Genome in a Bottle Consortium Data

To benchmark InDelible against GATK and Manta, we downloaded ES data for sample HG002 (Ashkenazi proband sample) provided by the Genome in a Bottle (GIAB) Consortium from the NCBI FTP site (https://github.com/genome-in-a-bottle/giab_data_indexes). We then acquired variant benchmarks for small variants from Zook et al. (2016)[16] and structural variants from Zook et al. (2020)[17] and limited each to variants with a REF/ALT size difference ≥1bp (i.e. InDels) and at least one breakpoint within padded (i.e. ±100bps) exome bait regions used for original whole exome sequencing (SureSelect coordinate files acquired from Agilent). We then ascertained variants from HG002 using InDelible, GATK, and Manta. For InDelible, we used the "complete" pipeline with default settings. Filtering was performed identically to that outlined for *de novo* variant discovery above, except we did not filter variants with high allele frequency, located outside coding sequence, or outside of known monoallelic DDG2P genes. For GATK we first ran HaplotypeCaller to generate a gVCF and then ran GenotypeGVCFs on the resulting output with default settings to generate a final output of genotyped sites. Default settings were used for GATK except we provided the SureSelect bait regions outlined above during the HaplotypeCaller step. We then used bcftools[18] to filter sites with GQ < 20 and DP < 7 to generate a final list of sites for benchmarking. To identify structural variant breakpoints with Manta, we first ran configManta.py with default settings other than providing ES baits with `--callRegions` and setting the `--exome` flag to generate a Manta workflow file. We then ran the resulting `runWorkflow.py` command.

We then converted all three callsets to bed format and used a custom python script to ask if any variant from HG002 as ascertained by the GIAB consortium was found within 100bps of a variant called by any of the three callers to calculate recall rate. All variants/breakpoints that were not within 100bps of a true variant were coded as false positives (Supplementary Figure 3). To calculate recall relative to InDelible, we quantified the number of variants >20bps in length recalled by InDelible, GATK, and Manta separately for deletions and insertions/duplications. We then divided calculated values for GATK and Manta by the value for InDelible as presented in the main text.

### Supplementary Figures

#### Supplementary Figure 1

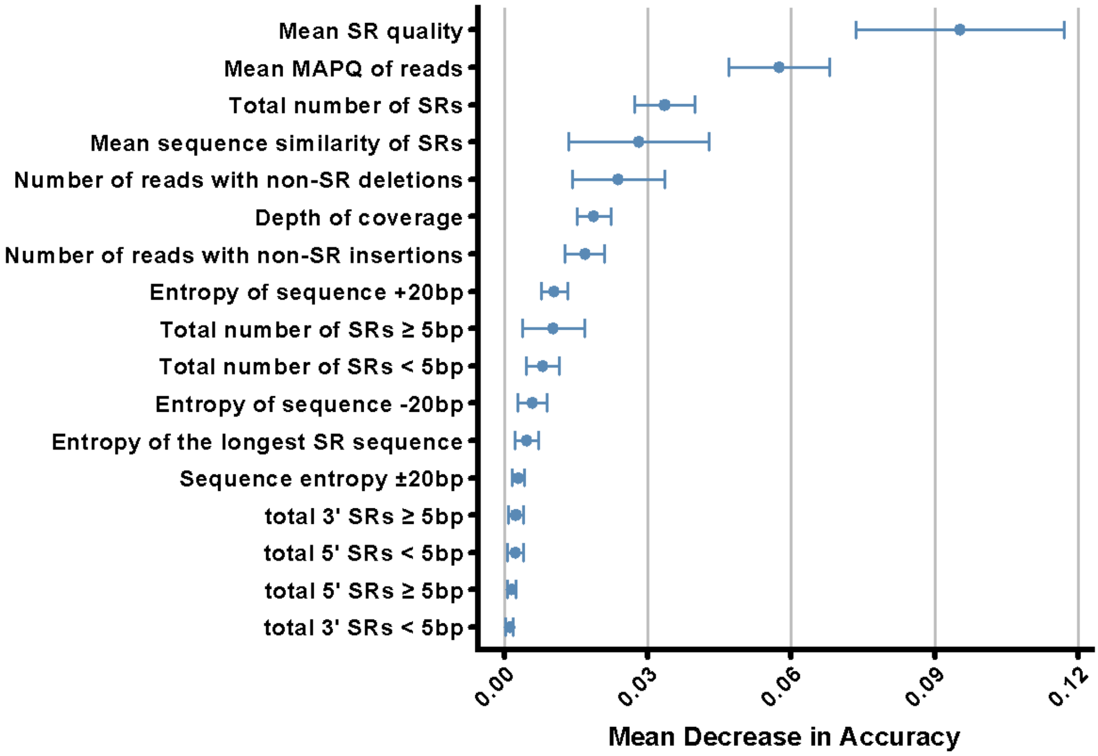


**Feature importance from the InDelible random forest.** Shown is the mean decrease in accuracy given by random forest for each of the 17 features we used to estimate the probability of a split read cluster being a true positive variant. Error bars represent standard deviation of mean decrease in accuracy for 10 cross-validated random forests at k = 75 (Supplementary Figure 4) for the final forest generated by our active learning model (see Supplementary Methods). Features which quantify the number of reads with clipped sequence length less than/greater than 5bp (e.g. Total number of SRs ≥ 5bp) represents the default setting of InDelible and can be adjusted by the end user during random forest training. Sequence entropy for all categories is calculated as in Schmitt and Herzel[1].

#### Supplementary Figure 2

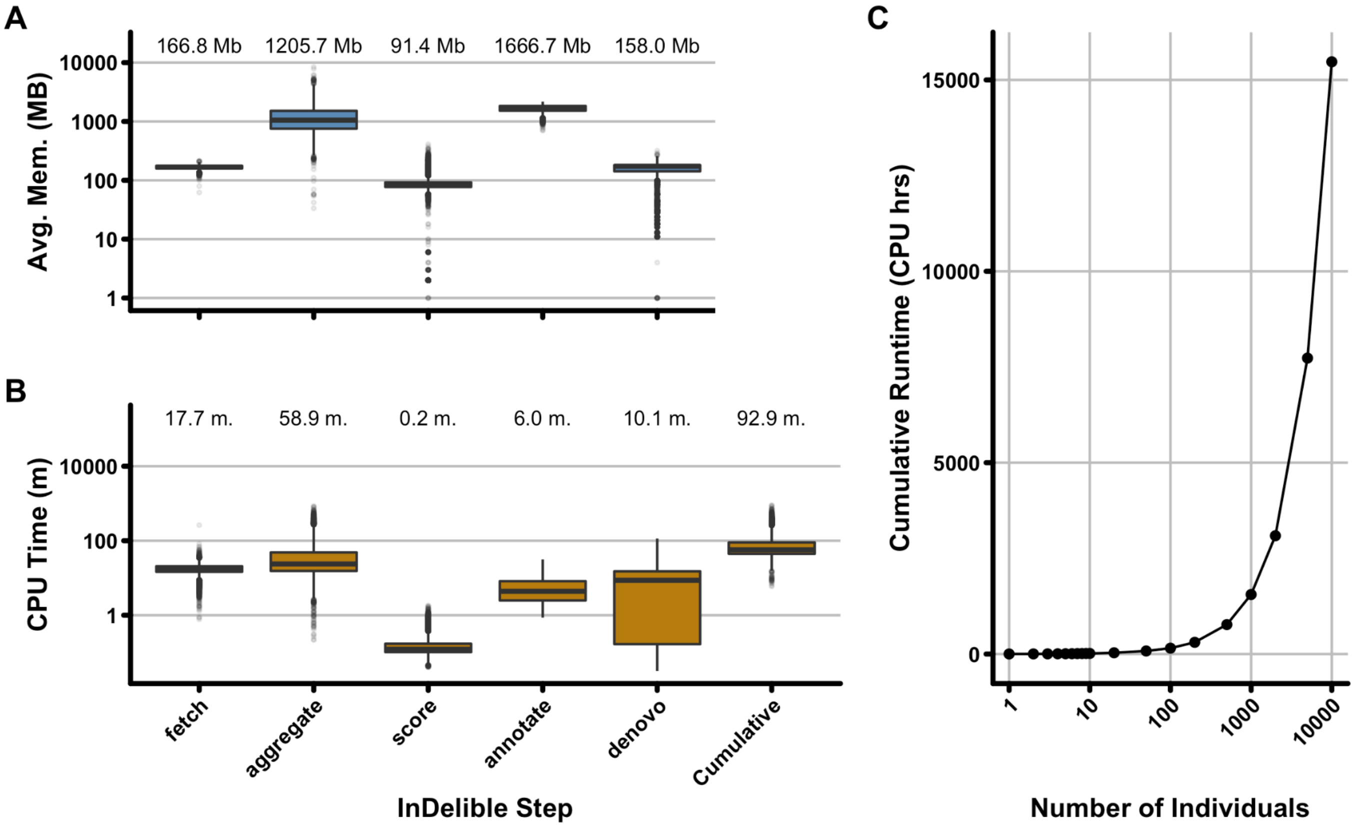


**Benchmarks of InDelible.** Using real data from the DDD study, we benchmarked the InDelible algorithm. Average memory use in megabytes (**A**) and CPU time in minutes (**B**) across all 13,438 DDD probands for the six primary steps of the InDelible SV discovery pipeline (Main Text Figure 1). Using the CPU times in (**B**), we extrapolated runtimes for studies of various sizes by randomly sampling the number of individuals shown on the x-axis (**C**).

#### Supplementary Figure 3

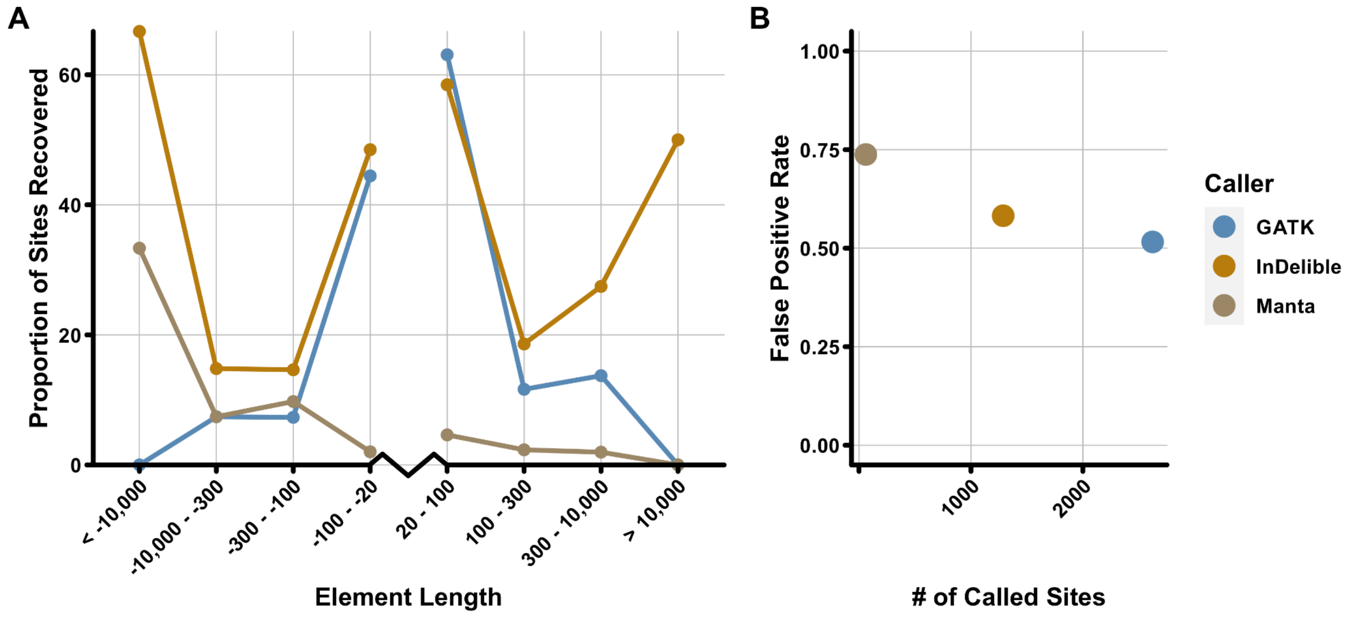


**InDelible benchmarks across the allele frequency spectrum.** To benchmark InDelible, we called variants using InDelible, GATK, and Manta in the genome of an Ashkanazi Jewish individual characterised for both small InDels and large structural variants by GIAB[16,17]. (**A**) Recall rate of all three callers for deletions (left side of plot) and insertions/duplications (right side of plot). Each point represents the proportion of variants in the size bin depicted on the x-axis. Size-ranges for deletions and insertions/duplications are right-open and left-open, respectively. Variants with an absolute size ≤20 base pairs were excluded from this analysis. Despite GATK generally being designed to identify variants <100bps in length, for a limited set of variants GATK was able to ascertain at least one breakpoint for some larger variants (i.e. > 100bps), albeit and as expected with incorrect size estimates. (**B**) False positive rate for all three callers as a function of total number of sites identified by each caller. We note that these experiments likely drastically overstate the FP rates of all callers – GIAB gold-standard calls were based on a merge of several different technologies and variants that are readily identifiable with long-read sequencing (e.g. Pacific Biosciences) may be very difficult to identify with short-read based approaches which query ES data.

#### Supplementary Figure 4

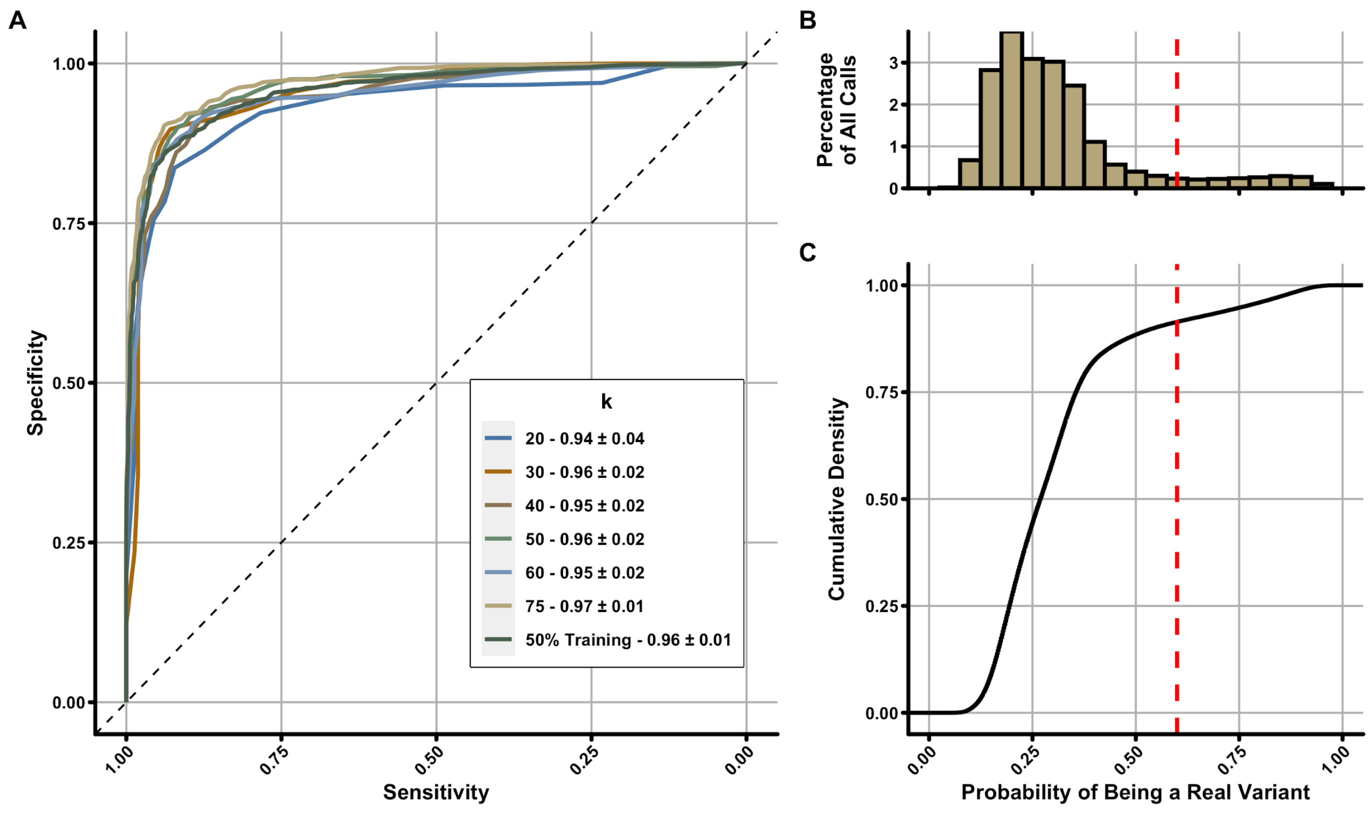


**Filtering split read clusters in InDelible with active learning.** To estimate the probability of a variant being real, we utilized an active learning approach (see Supplementary Methods). (**A**) Cross validated ROC curves at various inputs of *k* as well as for a traditional machine learning model where data was split into 50% training and test data without active learning. (**B**) Total number of original redundant split read clusters in 0.05 p-value bins. By default, InDelible filters all split read clusters with a probability of being a true variant <0.6 (red dashed line). (**C**) Cumulative density plot of the redundant clusters from (**B**).

#### Supplementary Figure 5

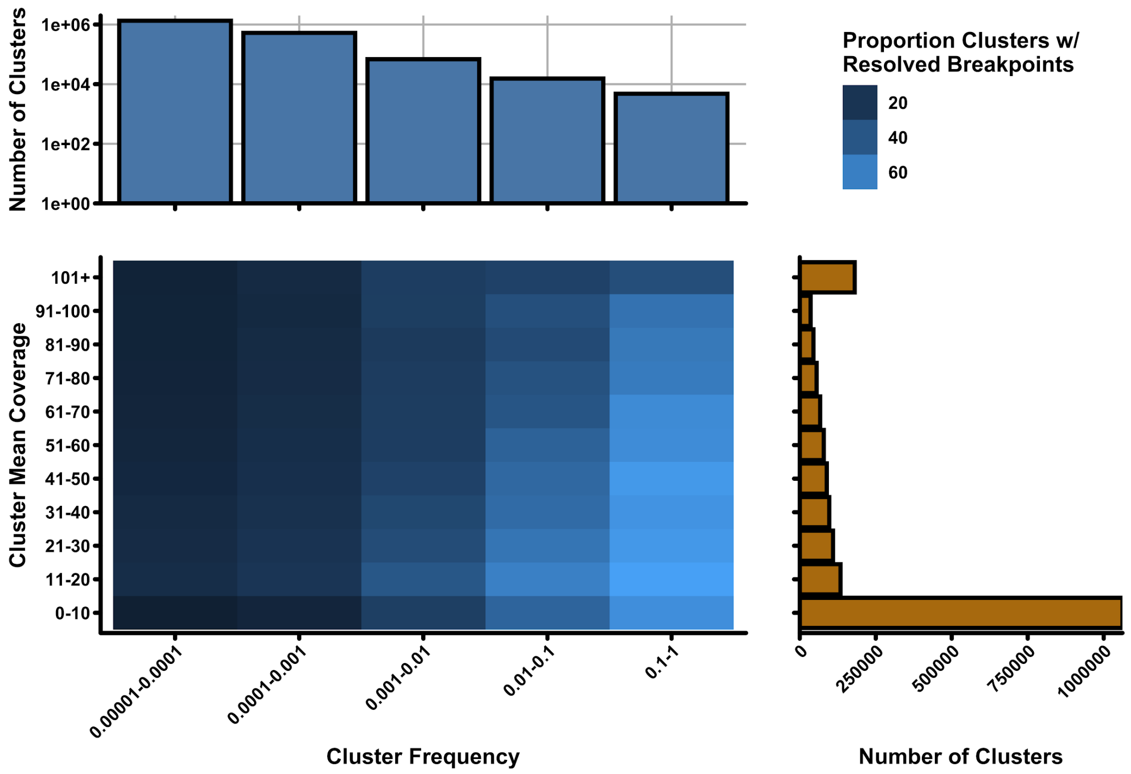


**Breakpoint resolution of InDelible as a factor of allele frequency and coverage.** Lower-left plot represents the proportion of sites within each bin on the X (cluster frequency – proportion of individuals a given site has been identified in) and Y (cluster mean coverage – mean sequencing coverage within individuals with a given cluster) axes which have resolved variant type and/or breakpoints. The lighter the shade of blue, the higher proportion of sites within that bin that have resolved breakpoints. Marginal histograms represent the total number of clusters within each bin on the X and Y-axes.

#### Supplementary Figure 6

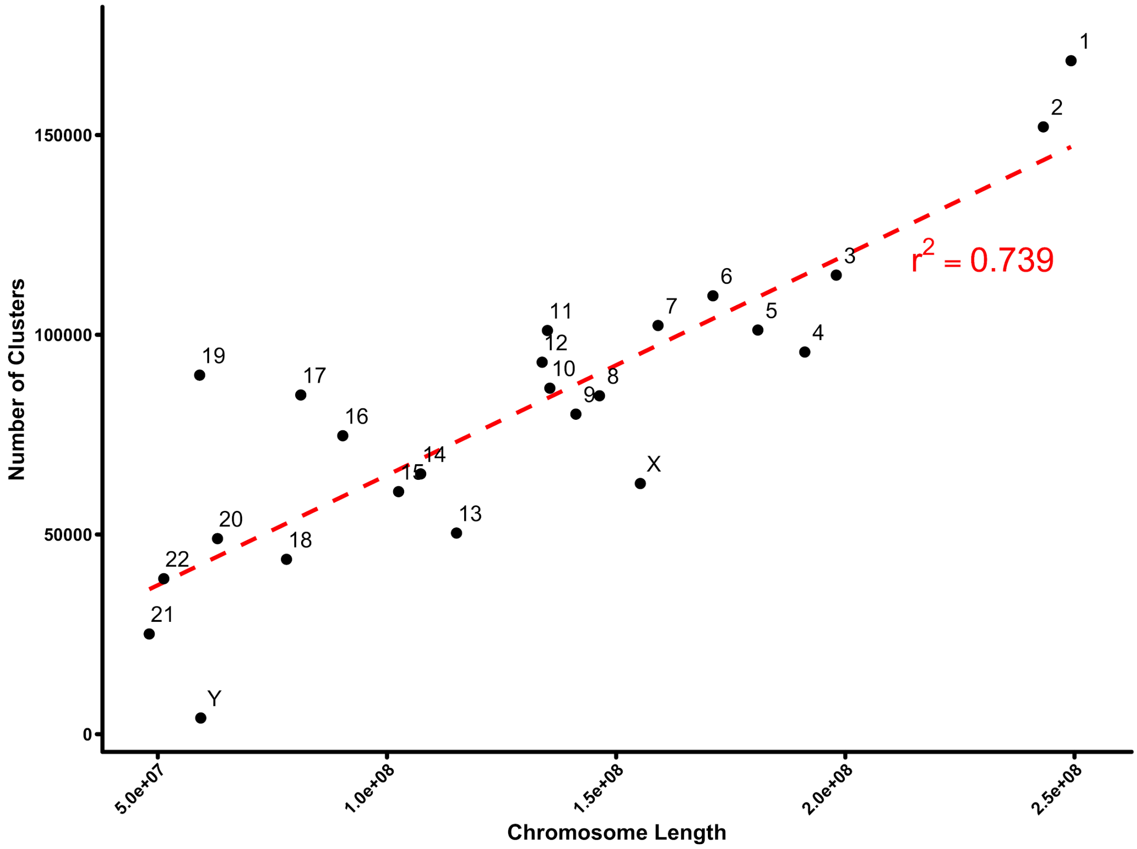


**Number of identified breakpoints per-chromosome.** Total number of breakpoint clusters identified on the 22 autosomes and 2 allosomes in the Deciphering Developmental Disorders study. The $r^2$ value represents the correlation coefficient for Number of Clusters ~ Chromosome Length.

#### Supplementary Figure 7

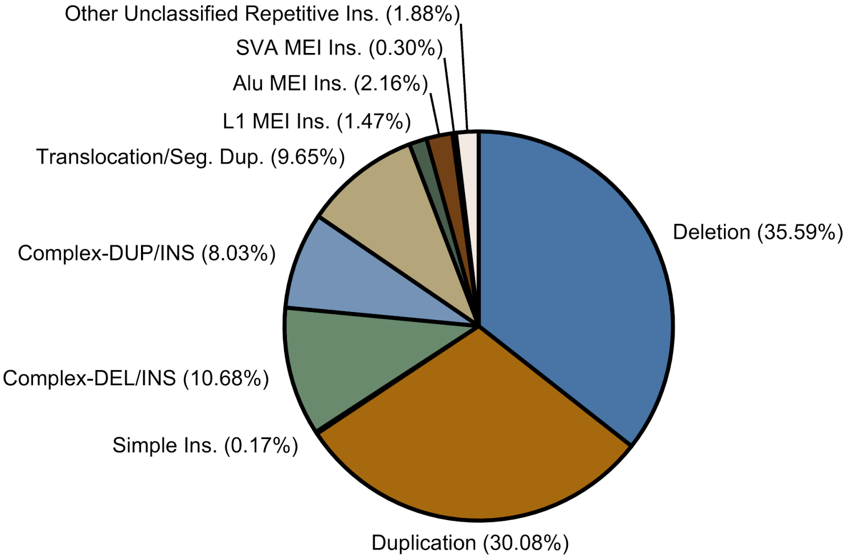


**Variant types identified by InDelible.** InDelible identifies a wide range of genetic rearrangement classes using a re-alignment of split read sequences to the reference genome. Each slice of the pie represents the proportion of breakpoints exhibiting the annotated structure among sites that have both 5'/3' breakpoints resolved or align to a known human repeat element (total n = 199,932 breakpoints). Translocations and segmental duplications (Translocation/Seg. Dup. above) are grouped together as discerning between these variant types is difficult with available sequencing data.

#### Supplementary Figure 8

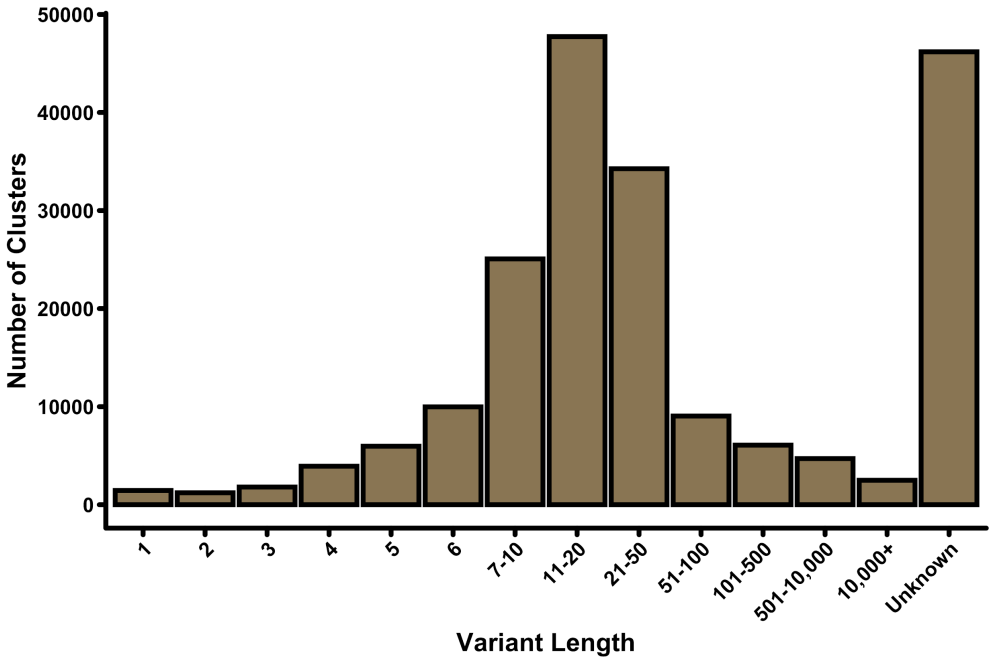


**Variants by resolved length.** Variants InDelible is able to generate length estimates for mostly fall between 10-100 base pairs in length. Variants with “Unknown” length are typically translocations, segmental duplications, or mobile element insertions. InDelible does not attempt to generate length estimates for these variant classes.

#### Supplementary Figure 9

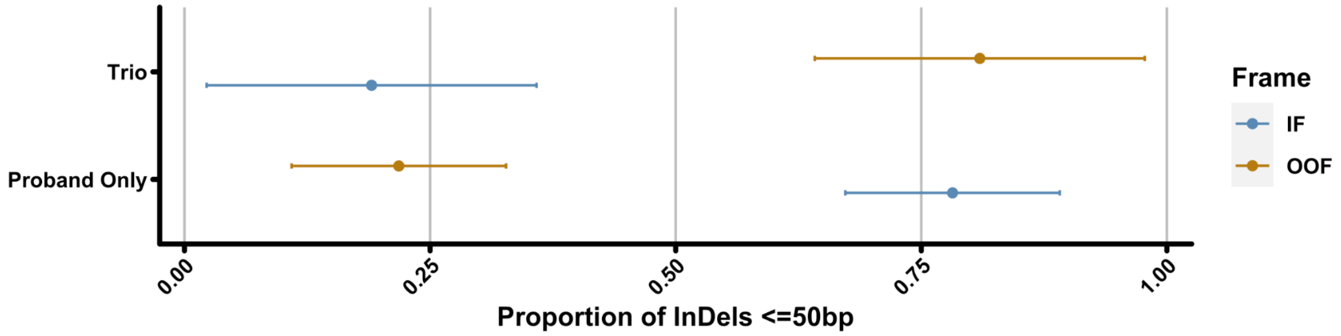


**Proportion of in-frame versus out-of-frame insertions for trio versus non-trio data.** Shown is the proportion of variants ≤50bp which are in-frame (blue) or out-of-frame (orange) separated by whether the variant was called in a proband sequenced with/without parental samples.

### Supplementary Tables

#### Supplementary Table 1

| Library Name | Version | Website | Citation (If Applicable) |
| --- | --- | --- | --- |
| Cython | 0.29.13 | https://cython.org/ | Behnel, et al.[19] |
| PyYAML | 5.1.2 | https://pyyaml.org/ | n.a. |
| Biopython | 1.74 | https://biopython.org/ | Cock, et al.[4] |
| Intervaltree | 3.0.2 | https://github.com/chaimleib/intervaltree | n.a. |
| numpy | 1.17.2 | https://numpy.org/ | Harris, et al.[20] |
| pandas | 0.25.1 | https://pandas.pydata.org/ | McKinney[21] |
| pybedtools | 0.8.0 | https://daler.github.io/pybedtools/ | Quinlan[22]; Dale, et al.[23] |
| pyfaidx | 0.5.5.2 | https://pythonhosted.org/pyfaidx/ | Shirley, et al.[24] |
| pysam | 0.15.3 | https://github.com/pysam-developers/pysam | n.a. |
| scikit-learn | 0.21.3 | https://scikit-learn.org/stable/ | Pedregosa, et al.[2] |
| scipy | 1.3.1 | https://www.scipy.org/ | Virtanen, et al.[25] |

**Python libraries used by InDelible.** “Version” reflects the version number of the named package used during software development and for all analyses performed in this manuscript.
